# Cross-sectional IgM and IgG profiles in SARS-CoV-2 infection

**DOI:** 10.1101/2020.05.10.20097535

**Authors:** Tugba Ozturk, J Christina Howell, Karima Benameur, Richard Ramonell, Kevin S. Cashman, Shama Pirmohammed, Leda C. Bassit, John D. Roback, Vincent C. Marconi, Raymond F. Schinazi, Whitney Wharton, F. Eun-Hyung Lee, William T Hu

## Abstract

**Background:** Accurate serological assays can improve the early diagnosis of severe acute respiratory syndrome coronavirus 2 (SARS-CoV-2) infection, but few studies have compared performance characteristics between assays in symptomatic and recovered patients.

**Methods:** We recruited 32 patients who had 2019 coronavirus disease (COVID-19; 18 hospitalized and actively symptomatic, 14 recovered mild cases), and measured levels of IgM (against the full-length S1 or the highly homologous SARS-CoV E protein) and IgG (against S1 receptor binding domain [RBD]). We performed the same analysis in 103 pre-2020 healthy adult control (HC) participants and 13 participants who had negative molecular testing for SARS-CoV-2.

**Results:** Anti-S1-RBD IgG levels were very elevated within days of symptom onset for hospitalized patients (median 2.04 optical density [OD], vs. 0.12 in HC). People who recovered from milder COVID-19 only reached similar IgG levels 28 days after symptom onset. IgM levels were elevated early in both groups (median 1.91 and 2.12 vs. 1.14 OD in HC for anti-S1 IgM, 2.23 and 2.26 vs 1.52 in HC for anti-E IgM), with downward trends in hospitalized cases having longer disease duration. The combination of the two IgM levels showed similar sensitivity for COVID-19 as IgG but greater specificity, and identified 4/10 people (vs. 3/10 by IgG) with prior symptoms and negative molecular testing to have had COVID-19.

**Conclusions:** Disease severity and timing both influence levels of IgM and IgG against SARS-CoV-2, with IgG better for early detection of severe cases but IgM more suited for early detection of milder cases.

## Introduction

The 2019 novel coronavirus disease (COVID-19) pandemic began in December 2019,^1,2^ and over 3 million people around the world have contracted the disease as of May 2020. Among both symptomatic and asymptomatic individuals with SARS-CoV-2, real time reverse-transcriptase polymerase chain reaction (rRT-PCR) remains the major confirmatory test. In the U.S., widespread rRT-PCR testing remains limited despite improvements. Moreover, rRT-PCR testing among clinical COVID-19 patients in China showed suboptimal sensitivity (positive in 72 of 104 sputum, 5 of 8 nasal swabs, 126 of 392 pharyngeal swabs).^3^ This is in keeping with previously identified challenges in the molecular diagnosis of the related SARS-CoV, including low viral count at onset, insufficient autopsy or neutralization tests as gold standard, and non-identical genetic strains.^4,5^ Several serological tests have been developed to detect immunoglobulins (IgG & IgM) against viral proteins,^6,7^ but serological tests face usual challenges of delayed positivity,^5^ host immune function^8^ and cross-reactivity to other coronaviruses.^9,10^ Design of epidemiological surveys and treatment trials can therefore be greatly hindered by the absence of a consensus laboratory diagnostic algorithm.

Similar to other coronaviruses, SARS-CoV-2 is composed of four structures: envelope, membrane, nucleocapsid, and spike.^2,11-13^ The majority of amino acids unique to SARS-CoV-2 are located in the receptor binding domain (RBD) of the S1 subunit,^14^ and S1 as well as the RBD domain have been used in serological assays for COVID-19.^6^ Previous work on SARS-CoV found increased envelope (E) protein levels during viral replication,^15^ and E proteins from the two beta coronaviruses only differ by four amino acids.^2^ S1 and E are therefore reasonable antigenic targets for serological assay development. Herein, we performed novel IgM (against the full-length SARS-CoV-2 S1 and highly homologous SARS-CoV E protein) assays and a commercially available IgG (against the S1 -RBD) assay in hospitalized and recovered COVID-19 patients, and compared their serological profiles with pre-2020 healthy control (HC) participants and people with negative SARS-CoV-2 rRT-PCR results (previously symptomatic or never-symptomatic).

## Materials and Methods

### Standard Protocol Approvals, Registrations, and Patient Consents

This study was approved by Emory University Institutional Review Board. Written consents were obtained from all participants or their legally authorized representatives (when appropriate).

### Study Participants

Four groups of subjects were included in the study: 1) **Hospitalized** symptomatic patients with moderate-to-severe influenza-like illness (ILI) in keeping with COVID-19 confirmed by rRT-PCR (n=18, with 14 requiring artificial ventilation; samples collected during hospitalization a median of 10.5 days after symptom-onset, range 4-24 days); 2) people who recovered from **mild** self-limited COVID-19 (n=14; nine with (+)rRT-PCR, four with ILI following direct contact with confirmed COVID-19 cases but not eligible for rRT-PCR, and one with ILI following direct contact with confirmed COVID-19 cases but did not seek rRT-PCR; samples collected a median of 18.5 days after initial symptom onset, range 9-33); 3) **pre-2020 HC** (n=103) recruited through inflammation studies targeting the young (PI: WTH),^16^ middle-aged (PI: WW),^17^ or older (PI: WTH) adults; and 4) people who had **(-)rRT-PCR** results in 2020 (n=13; two symptomatic at time of draw, eight recovered from mild self-limited ILI, and three never had any symptoms; none had follow-up rRT-PCR). Sample size was calculated based on one previous study^6^ when the current study began using a more conservative effect size (0.8 vs. >1), with an estimated disease prevalence of 5%-20%. Plasma was collected from five hospitalized participants, nine mild participants, and all pre-2020 HC and those with (-)rRT-PCR. Serum was collected from the remaining 13 hospitalized participants and five mild participants.

### Serological Assays

A commercial anti-S1 receptor binding domain (RBD) IgG indirect ELISA assay (GenScript, Piscataway, NJ) was purchased and performed per manufacturer’s protocol, except two plasma dilutions (1:16 and 1:64) were selected from a range of 1:8 – 1:256 performed in a subgroup of COVID-19 and pre-2020 HC subjects.

For IgM, we developed two novel assays. Synthetic SARS-CoV-2 S1 (230-01101-100, produced from *E. coli)* and SARS-related E (228-11400-2, produced from *E. coli)* proteins were purchased from RayBiotech (Peachtree Corners, GA). For IgM, 100 μL of 2.5 μg/mL antigen in PBS was applied to standard 96-well plate at 4°C overnight. Six (out of 96) wells were coated only with 5% albumin without S1/E. Plates were washed with PBS before blocking at room temperature for 1 hr with 4% non-fat dried milk (nfdm). Diluted plasma samples (1:64, 1:64, 1:256, 1:1024 in PBS containing 2% nfdm and 0.1% Tween20) were loaded into blocked wells for 1 hr. Wells were then washed three times with PBS, and 50 μL of 1:20,000 goat anti-human IgM fc (09-035-043, Jackson ImmunoResearch Laboratories, West Grove, PA) was added to each dilution condition for 30 min. Wells were treated with strepavidin-HRP (1:200, 50 μL per well) for 20 min in the dark, washed, incubated with substrate mix for 20 min in the dark, and treated with reaction stop solution. Plates were then read at 450 nm (Molecular Devices, SpectraMax-M2) followed by background (570 nm) subtraction.

### Statistical Analyses

All statistical analyses were performed using SPSS 26 (IBM SPSS, Armonk, NY) except for curve-fitting. Differences in optical densities (OD) were calculated at 1:16 dilution for the commercial IgG assay and 1:128 dilutions for all IgM assays. Chi-squared or Fisher’s exact test was used to analyze differences between symptomatic and recovered COVID-19 patients, and Student’s T-test was used to analyze differences between these two groups’ age, anti-S1 and anti-E IgM levels, and log_10_-transformed anti-S1-RBD IgG due to its non-normal distribution. Duration of disease was available for 8/14 (57%) of mild patients, and only available values were used for descriptive analysis.

Curve-fitting for relationships between antibody levels and time since symptom onset was performed in GraphPad Prism 8.4.2 (San Diego, CA). For each antibody, linear regression was compared against other higher order models (second- or third-order polynomial, and exponential growth for anti-S1-SBD IgG in recovered cases) based on Akaike Information Criteria. Except for anti-S1 IgM in hospitalized participants, linear functions provided better fit than more complex models.

Receiver-operating characteristic (ROC) curve analysis was first used to determine each serological test’s ability to distinguish between symptomatic COVID-19 cases and 78 randomly selected pre-2020 HC. Threshold values from these ROC curve analyses were tested in the recovered cohort against 25 pre-2020 HC subjects. Given differences in the symptomatic and recovered groups, we further performed 100-fold ROC curve analysis using either anti-S1-RBD IgG or the product of anti-S1 and anti-E IgM. For each run, COVID-19 cases were randomly assigned to the training or test set at 1:1 ratio, and pre-2020 HC cases were randomly assigned to the training or test set at 2:1 ratio. Thresholds were automatically determined in the training set to maximize accuracy while maintaining balance between sensitivity and specificity, and applied to the test set to determine outcome sensitivity and specificity. Median threshold values from the 100-fold ROC curve analysis were used in the group of people with negative molecular testing.

Given the expected effect sizes, Bonferroni correction was used to adjust for multiple comparisons.

## Results

Compared to hospitalized cases, mild cases were younger (median age 31.5 vs. 61.5 years, t(28)=3.593, p=0.001) and did not have any African Americans (0 vs. 72%, p<0.001). Compared to pre-2020 HC participants, hospitalized cases had greater anti-S1-RBD IgG (log_10_ transformed due to non-normal distribution, t(31.7)=10.816, p<0.001, Fig 1a), anti-S1 IgM (t(119)=5.129, p<0.001, Fig 1b), and anti-E IgM (t(119)=4.121, p<0.001, Fig 1c). The same was true among mild cases for anti-S1-RBD IgG (t(115)=4.042, p=0.001, Fig 1a), anti-S1 IgM (t(117)=6.967, p<0.001, Fig 1b), and anti-E IgM (t(115)=3.872, p<0.001, Fig 1c) compared to pre-2020 HC cases. Regression analysis showed women to have higher anti-S1 (F(1,128)=6.22, p=0.014) and anti-E (F(1, 128)=7.08, p=0.009) IgM levels independent of COVID-19 status, but sex did not influence IgG levels.

**Figure 1.**
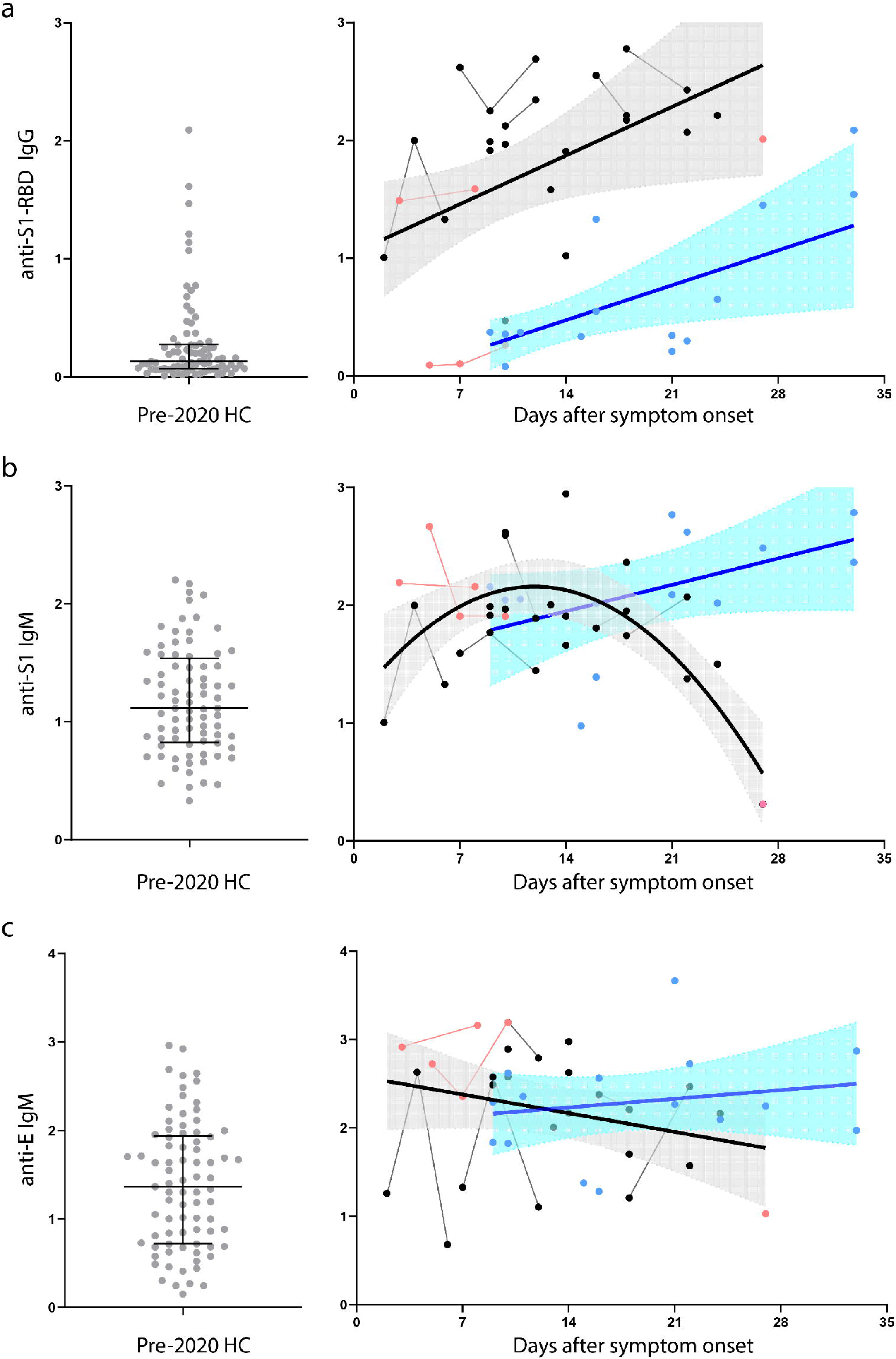
Serological assay results of COVID-19 participants. Anti-S1-RBD IgG (a), anti-S1 IgM (b), and anti-E IgM (c) levels were analyzed in pre-2020 HC participants (gray circles), hospitalized symptomatic COVID-19 participants with severe (black circles) or mild-to-moderate (red circles) disease, and COVID-19 participants who had recovered from mild self-limited disease (blue circles). Antibody levels for COVID-19 were plotted according to self-reported symptom onset. Thin lines between represent serial sampling from the same subject, and thick lines with 95% confidence intervals represent best fit lines.

Using these hospitalized cases and 78 pre-2020 HC, ROC analysis showed anti-S1 IgG to have AUC of 0.942 (95% CI 0.883-1.000; Fig 2a), with a cut-off of 0.89 OD associated with 88.9% sensitivity and 92.3% specificity. However, this cut-off identified only four of 14 (28.6%) mild participants, with the mild cases having much lower levels than hospitalized cases (0.71 vs. 1.77, t(30)=4.261, p=0.0002, Fig 1a). Linear regression analysis (which better fit data than higher order polynomials) of the cross-sectional cohorts showed the mild cases’ IgG levels to rise at the same rate (slope 0.042, 95% CI: 0.009-0.075) as in the hospitalized cohort (slope 0.059, 95% CI: 0.008-0.110) but lagged the latter by 28 days. In contrast, neither anti-S1 IgM (t(30)=1.703, p=0.099) nor anti-E IgM (t(30)=0.190, p=0.850) differed between mild and hospitalized cases, although IgM levels had a downward trend with longer disease duration in the hospitalized cases. Extrapolating the linear IgG (OD=1.04+0.059*days) and the second-order anti-S1 IgM (OD=2.16-0.00348 * (days-12.2)-0.00699*(days-12.2)^2^) curves among hospitalized participants showed a pre-symptomatic incubation period of 16 days vs. 5.6 days.

**Figure 2.**
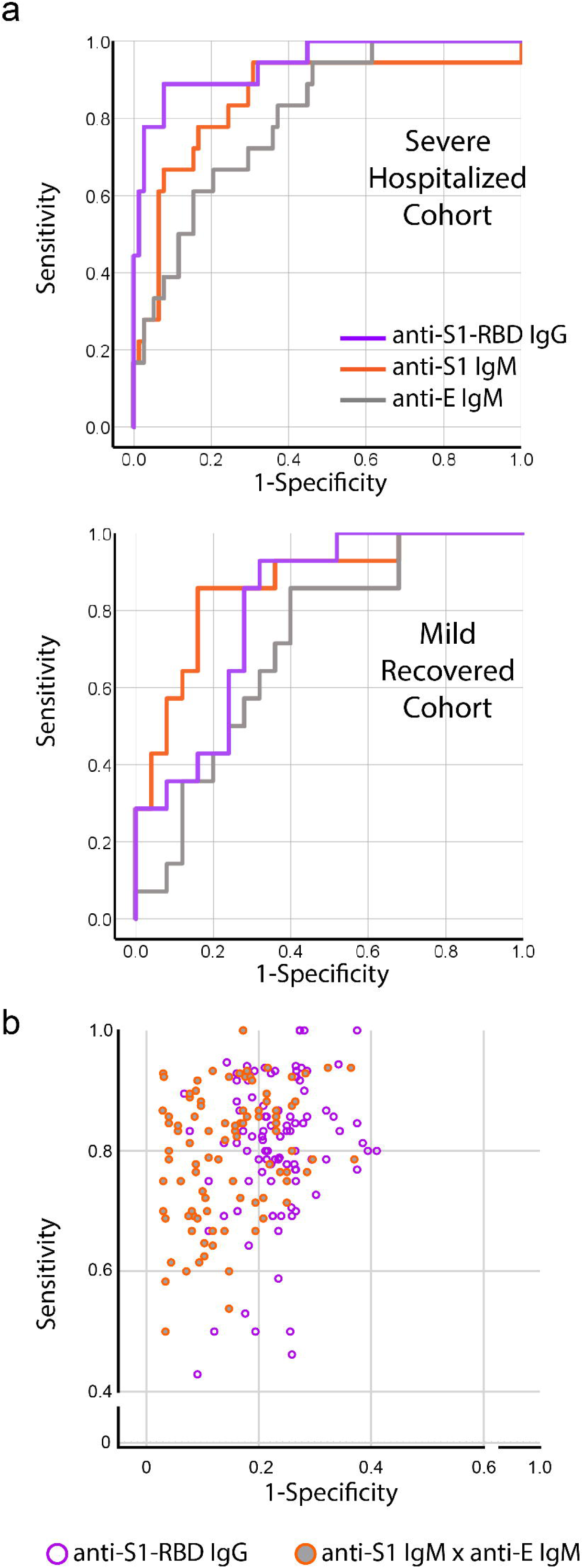
Receiver operating characteristics curve analysis showing performance differences between hospitalized and mild participants (a). 100-fold ROC curve analysis showed similar sensitivity between anti-S1-RBD IgG and the combination IgM (product of anti-S1 and anti-E IgM), but the latter has greater specificity (p<0.0001).

ROC analysis of anti-S1 IgM (AUC=0.852, 95% CI of 0.739-0.965) with a cut-off of 1.37 OD was associated with sensitivity of 94.4%, specificity of 69.2%, and detection of 13/14 (92.8%) mild participants; anti-E IgM (AUC=0.807, 95% CI of 0.707-0.907) with a cut-off of 2.00 OD was associated with sensitivity of 66.7%, specificity of 79.5%, and detection of 9/14 (64.2%) mild participants. Because anti-S1 IgM is more sensitive while anti-E IgM is more specific, we multiplied the two IgM levels to achieve a balance between sensitivity and specificity (Table 2).

**Table 1.** Demographic and other information included in the current study. Categorical and continuous variables which differed between groups are shown in bold. * Comparison between symptomatic, recovered, and (-)rRT-PCR groups only. † Different between hospitalized and mild cases at p<0.005. †† n=8 for mild and n=10 for (-)rRT-PCR.

|  | Hospitalized<br>n=18 | Mild<br>n=14 | Pre-2020 HC<br>n=103 | (-)rRT-PCR<br>n=13 | p |
| --- | --- | --- | --- | --- | --- |
| Female (%) | 7 (39%) | 6 (43%) | 61 (59.2%) | 9 (69%) | 0.434 |
| **Age, median (range), yr** | **61.5†**<br>**(30-81)** | **31.5†**<br>**(26-81)** | **62**<br>**(24-87)** | **35.8**<br>**(29-63)** | **<0.0001** |
| Race |  |  |  |  | **<0.0001** |
| Asian | 2 (11%) | 0 | 2 (2%) | 1 (8%) |  |
| **African American** | **13 (72%)†** | **0†** | **15 (14%)** | **3 (23%)** |  |
| Non-Hispanic Caucasian | 3 (17%) | 12 (86%) | 82 (80%) | 8 (61%) |  |
| Hispanic | 0 | 1 (7%) | 0 | 1 (8%) |  |
| Other | 0 | 1 (7%) | 4 (4%) | 0 |  |
| **Inpatient/outpatient\*** | **18/0** | **1/13** | **-** | **0/13** | **<0.001** |
| Days since symptom onset,<br>median (range)* | 10.5<br>(4-24) | 18.5<br>(9-33) | - | 17<br>(2-43) | 0.057* |
| **Respiratory failure**<br>**requiring intubation (%)** | **14 (78%)** | **0** | **-** | **0** | **<0.001** |
| Duration of disease, median<br>(range), days | 10.5<br>(4-24) | 11<br>(4-23)† |  | 4.5<br>(1-30) | 0.126 |
| Clinical symptoms* |  |  | - |  |  |
| Cough | 16 (89%) | 9 (64.3%) | - | 6 (46%) | 0.036 |
| Fever/chills | 15 (83%) | 10 (71%) | - | 5 (38%) | 0.030 |
| Shortness of breath | 13 (72%) | 6 (43%) | - | 3 (23%) | 0.022 |
| Myalgia | 6 (33%) | 9 (64%) | - | 4 (31%) | 0.130 |
| Headaches | 5 (28%) | 8 (57%) | - | 2 (15%) | 0.058 |
| Sore throat | 2 (11%) | 6 (43%) | - | 5 (38%) | 0.096 |
| Nasal congestion/rhinorrhea | 2 (11%) | 6 (43%) | - | 4 (31%) | 0.121 |
| Diarrhea | 2 (11%) | 3 (21%) | - | 3 (23%) | 0.630 |
| **Anosmia** | **0<sup>†</sup>** | **6 (43%)<sup>†</sup>** | **-** | **0** | **<0.001** |
| Fatigue | 1 (6%) | 2 (14%) | - | 2 (15%) | 0.623 |
| Vomiting | 0 | 1 (7%) | - | 0 | 0.323 |
| Never symptomatic* | 0 | 0 | - | 3 (23%) | 0.021 |
| **SARS-CoV-2 detected by**<br>**rRT-PCR\*** | **16/16** | **9/9** | **-** | **0/13** | **<0.001** |
| **Anti-S1-RBD IgG, median**<br>**(range), OD** | **2.04<sup>†</sup>**<br>**(0.15-2.73)** | **0.37<sup>†</sup>**<br>**(0.08-2.09)** | **0.12**<br>**(0.01-2.09)** | **0.28**<br>**(0.02-0.91)** | **<0.001** |
| **Anti-S1 IgM, median (range)**<br>**OD** | **1.91**<br>**(0.31-2.95)** | **2.12**<br>**(0.98-2.78)** | **1.14**<br>**(0.33-2.53)** | **2.07**<br>**(0.25-3.36)** | **<0.001** |
| **Anti-E IgM, median (range),**<br>**OD** | **2.23**<br>**(1.03-3.04)** | **2.26**<br>**(1.28-3.67)** | **1.52**<br>**(0.15-3.04)** | **2.58**<br>**(0.46-3.10)** | **<0.001** |

**Table 2.**
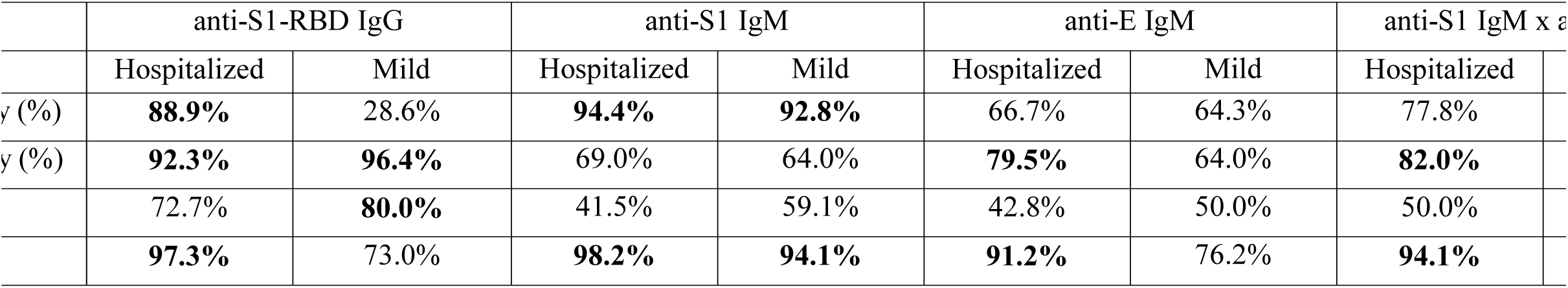
Performance characteristics of three serological tests in the hospitalized (training, vs. 78 pre-2020 HC) and mild (validation, vs. 25 pre-2020 HC) cohorts, using thresholds developed in the hospitalized cohorts.

As an alternative to using a training cohort consisting of entirely hospitalized cases, we performed 100-fold simulation using a training cohort of randomly selected COVID-19 and pre-2020 HC participants (1:1 and 2:1 distribution between the training and test groups, Fig 2b). This analysis showed – in the test groups – a median sensitivity and specificity of 82.1% and 86.0% for anti-S1 x anti-E IgM, vs. 82.4% and 76.5% for anti-S1-RBD IgG. The combined IgM was more specific than IgG (t(189.95)=8.393, p<0.0001), but the two assays had similar sensitivity (t(198)=0.669, p=0.504). Median thresholds of 3.58 OD^2^ and 0.31 OD identified seven and six of the 13 (-) rRT-PCR participants as having positive serology, with four having elevated levels of both. Taking into account days from symptom onset among those who reported ILI symptoms (n=10), four had elevated IgM product and three of the four also had elevated IgG levels.

## Discussion

The diagnosis and sero-surveillance of COVID-19 can be challenging. While small numbers of (+)rRT-PCR patients had high antibody titers against the S1 protein,^6^ patients who recovered – the focus of any sero-survey – have not been systemically investigated. Here we show that a serological assay with reactivity against the most commonly targeted antigen (S1-RBD) had high sensitivity and specificity among hospitalized participants, but its sensitivity dramatically diminished in participants with mild COVID-19 during the first month after symptom onset. The lower anti-S1-RBD IgG levels may be related to milder severity, shorter disease duration, or both. In contrast, IgM levels were more comparable between symptomatic and recovered patients, although only more severe cases showed lower IgM levels with greater disease duration. Because of the time-dependent variability in these antibody levels, the combined IgM levels had greater specificity than IgG in detecting hospitalized and mild participants without loss in sensitivity during the first month. Early IgM and IgG profiles should be tested for their predictive value in distinguishing between severe (elevated IgM and IgG) and mild (elevated IgM, normal IgG) outcomes in a prospective cohort.

A diagnostic algorithm using IgG levels trained on our hospitalized participants performed poorly to detect those who had mild COVID-19. This is generally in keeping with results from China showing low or medium-low neutralizing antibody titers in 47% of patients who recovered from mild COVID-19,^18^ although it is difficult to interpret whether the neutralizing antibodies identified represented IgM, IgG, or both. The slow rise in IgG levels has also been reported by the UK National COVID Testing Scientific Advisory Panel using a novel assay against the SARS-CoV-2 trimeric spike protein,^7^ and was previously observed in SARS-CoV cases.^19^ The longer persistence of IgM may then be a corollary of the slow IgG increases, with long-lived antigen-induced plasma cells^20^ implicated in similar IgM persistence in other viral^21,22^ and non-viral^23^ infections. Questions remain regarding whether the hospitalized severe cases have a long pre-symptomatic incubation period with slopes similar to their IgM and IgG profiles during the symptomatic phase, with the extrapolated incubation period (5.6 days) from the IgM curve more in keeping with current knowledge than the extrapolated value (14 days) from IgG. It also remains to be seen whether mild cases’ IgM and IgG profiles would follow those of severe cases months out from their symptomatic phase. Finally but importantly, the expected heterogeneity in anti-S1-RBD IgG within hospitalized and mild cases needs detailed investigation as it may account for differences in neutralization potential, total IgG levels, and disease severity.

Altogether, the two divergent temporal profiles of IgM and IgG suggested by our cross-sectional studies need further confirmation from longitudinal within-individual studies whose results will have significant implications in viral surveillance, post-exposure immunity, and vaccine development.

These studies also highlight the importance of harmonizing serological testing methods and findings among COVID-19 cohorts according to symptom onset and severity. Hospitalized cohorts are often used for assay development because they had the greatest access to rRT-PCR testing and, conversely, were most accessible to clinical researchers. The choice of serological test can therefore underestimate past exposure to SARS-CoV-2, over-estimate immunity for convalescent plasma,^24^ or influence the choice of point-of-care lateral flow assays in large sero-surveillance studies.^7^ Until a gold standard better than rRT-PCR is confirmed, rapid development of a standardized cohort (including clinically suspected COVID-19 with and without rRT-PCR confirmation of various severity at multiple time points) with adequate reference biofluid samples is urgently needed to empirically assess the performance of novel as well as marketed serological tests.

While our study included repeated samples only in a few individuals and the overall cohort size is limited, the broad cross-sectional inclusion both symptomatic and recovered patients provide an overview of IgM and IgG profiles in COVID-19 relative to time since symptom onset. The over-representation of African Americans in the more severely ill cohort may mediate some differences in antibody profiles.^25^ Further work is also necessary to determine antibody levels, if measured early in disease course, can adequately predict severity of disease. However, IgG clearly has a role in confirming severe COVID-19 cases, and a commercially available option such as the one we used can accelerate broad diagnostic testing independent of, or in addition to, single-center efforts which are more difficult to standardize. Levels of IgM and IgG – against multiple viral proteins or different configurations of the same protein – should also be routinely measured during *in vitro* neutralization experiments and convalescent plasma trials. Finally, because we found a complex relationship between antibody levels, disease severity, and time since symptom onset, we urge extreme caution in using point-of-care or a single serologic assay to inform public policies.

## Data Availability

Data available upon reasonable request one year following publication.

## Author contribution/Acknowledgments

TO, JCH, VCM, RFS, and WTH contributed conception and design of the study; TO, JCH, KB, RPR, KSC, LCB, VCM, JDR, WW, FEL, and WTH contributed to acquisition, analysis, and interpretation of the data; WTH performed the statistical analysis; TO, JCH, and WTH drafted the manuscript; KB, RPR, KSC, LCB, JDR, VCM, WW, and FEL provided critical revisions for important intellectual content; all authors read and approved the submitted version.

## Acknowledgements

This work was supported by National Institutes of Health grants R01 AG 054046, R01 AG054991, and T32HL116271.

